# Deep Learning Reconstruction for Ultra-High-Resolution Photon-Counting CT of the Lung: Image Quality and Texture Characterization

**DOI:** 10.64898/2026.09.04.26361840

**Authors:** Kai Mei, Leonid Roshkovan, Leening P. Liu, Sandra S. Halliburton, Shobhit Sharma, Steven Ross, Tyler Stroud, Naruomi Akino, Zhou Yu, Richard Thompson, Ali H. Dhanaliwala, Harold I. Litt, Peter B. Noël

## Abstract

**Background:** Deep learning reconstruction (DLR), in contrast to hybrid iterative reconstruction (IR), offers an alternative approach to mitigate noise and improve contrast in ultra-high-resolution (UHR) photon-counting CT (PCCT).

**Purpose:** To characterize image noise, lesion contrast, spatial resolution, and attenuation distribution for DLR using a patient-derived 3D-printed phantom in UHR PCCT, and to apply the same analytical framework to patient examinations acquired on the same system.

**Methods:** A patient CT scan-derived lung phantom containing three digitally inserted lesions of differing morphology was fabricated using *PixelPrint* 3D printing and imaged on a CZT-based PCCT prototype at four dose levels (CTDI_vol_ 6, 4, 2, and 1 mGy). Images were reconstructed with hybrid IR at normal resolution (512 × 512) and UHR (1024 × 1024), and with DLR at UHR. Image noise, lesion contrast, and contrast-to-noise ratio (CNR) were measured within prespecified regions of interest replicated across reconstructions by coordinate transfer. Spatial resolution was assessed by the modulation transfer function (MTF) from a high-contrast edge at eight locations. Attenuation distribution and local texture were characterized using histograms and gray-level co-occurrence matrices (GLCMs) within a structured parenchymal region. Two patient examinations with part solid lung nodules acquired on the same prototype were analyzed as illustrative examples, with solid and ground-glass components segmented separately and their attenuation distributions compared.

**Results:** In the phantom, DLR reduced image noise relative to UHR hybrid IR (7.8% at 6 mGy and 15.2% at 1 mGy). Lesion contrast was constant across reconstructions and dose levels, so CNR differences reflected differences in noise alone. MTF curves for DLR lay above those for hybrid IR across the evaluated spatial frequency range at both 6 and 1 mGy, averaged +0.4 lp/mm in MTF50. In a structured parenchymal region, the attenuation histogram mode became progressively narrower from hybrid IR at 1 mGy to DLR at 6 mGy (averaged −23%), while mode position and mean regional attenuation were preserved, consistent with reduced partial-volume mixing rather than altered attenuation. In the patient example, the attenuation distributions of solid and ground-glass components were more widely separated with UHR DLR (2.0) than with either hybrid IR(1.6).

**Conclusions:** In UHR photon-counting CT of the lung, DLR reduced image noise while preserving lesion contrast and regional attenuation, increased measured MTF at a high-contrast edge, and produced attenuation distributions consistent with reduced partial-volume mixing at material boundaries. The same effect was observed in the separation of solid and ground-glass components in two patient examples.

## Introduction

Computed tomography is central to the diagnosis and management of lung disease. Many clinically consequential findings, however, occur at spatial scales that approach the resolution limits of current clinical CT: the internal structure and accurate delineation of subsolid nodule margins, early fibrotic change, small-airway wall thickening, and subtle emphysema. At these scales, image noise, partial-volume averaging, and reconstruction-dependent texture together determine whether a finding is visible at all and whether it can be measured reproducibly.

Photon-counting CT (PCCT) records individual photons and their energies rather than integrating deposited charge over time. This removes electronic noise from the measurement and permits smaller detector elements without the geometric dose penalty imposed by anti-light spreading septa in energy-integrating designs, thereby supporting ultra-high-resolution (UHR) acquisition [1], [2], [3], [4]. For thoracic imaging these properties are attractive because the lung is dominated by high-contrast interfaces, e.g., airway walls, small vessels, interlobular septa, whose depiction is resolution-limited rather than contrast-limited [5], [6], [7]. UHR acquisition, however, is not free of consequences: smaller detector elements collect fewer photons per element, so at fixed dose the noise magnitude increases, and the noise spectrum shifts toward higher spatial frequencies. The penalty is greatest at the low doses used for routine and screening chest CT [8], [9].

Deep learning reconstruction (DLR) offers a software route to suppress that noise. Hybrid iterative reconstruction (IR) reduces noise magnitude but can shift noise power toward lower spatial frequencies, producing the artificial, blotchy appearance widely described in the literature, and can smooth low-contrast structure in a spatially varying way [10]. DLR is instead trained to reduce noise while preserving edge fidelity and image texture [11], [12], [13], [14]. The DLR network learns is nonlinear and object-dependent, which has two consequences that are central to this work. First, its behavior cannot be extrapolated from measurements made on simple geometric phantoms, because performance depends on the object being imaged. Second, denoising that is aggressive relative to the information present in the data can suppress genuine low-contrast structures or, in the opposite, produce structure that is not supported by the measurement. In the lung, where the findings of interest are fine reticular structures, both failure modes are clinically consequential, and neither is detectable by inspection of patient images alone.

Previous clinical studies have demonstrated the potential of UHR PCCT for lung imaging [15], [16], [17], [18]. Several groups have characterized reconstruction settings for UHR photon-counting detector CT of the lung, focusing on IR strength, convolution kernel, and slice thickness [19], [20], [21]. On a CZT-based photon-counting detector CT platform, Sasaki and colleagues [19] compared normal-resolution and UHR hybrid IR with UHR DLR in 25 patients with lung nodules and reported reader-assessed improvements in image quality and diagnostic confidence. That work establishes perceived benefit in patients, but reader assessment *in vivo* cannot separate preserved anatomical structure from generated texture, because no ground truth is available. Conversely, our group has previously characterized a commercial DLR algorithm against patient-derived 3D-printed phantoms across a wide dose range, but on an energy-integrating platform at conventional resolution [22]. The behavior of DLR in the UHR PCCT regime specifically, where noise magnitude is higher, its spectral distribution differs, and the reconstructed voxel approaches the scale of the structures of interest, has not been characterized against a known reference.

The purpose of this study was therefore twofold. First, to characterize image noise, lesion contrast, spatial resolution, and local image texture for DLR relative to hybrid IR in UHR PCCT of the lung, using a patient-derived 3D-printed phantom in which the underlying structure is known. Second, determine whether a texture measure defined under those controlled conditions can be applied unchanged to patient images acquired on the same system, thereby providing a common quantitative operator linking phantom measurements to clinical image appearance.

## Methods

### Phantom Design and Fabrication

A lifelike lung phantom was fabricated using *PixelPrint* 3D-printing technology, which converts patient DICOM data directly into printer instructions and modulates the ratio of extruded filament to voxel volume so that each voxel reproduces a prescribed attenuation [23], [24]. Source imaging data used to generate the phantom were derived from an ultra-high-resolution CT dataset acquired on a Canon Aquilion Precision CT scanner (Canon Inc., Otawara, Japan).

Three digital lesions were inserted into the lung volume prior to printing [25], [26]. Lesion 1 was a highly spiculated spherical lesion, 27 mm in diameter. Lesion 2 was a moderately spiculated spherical lesion, 13 mm in diameter. Lesion 3 was a mildly spiculated ovoid lesion measuring 10 mm in plane. The phantom was produced by fused deposition modeling on a 3D printer (Original Prusa XL, Prusa, Prague, Czech Republic) fitted with a 0.25 mm brass nozzle. PLA filament (Atomic Filament, Kendallville, IN, USA) of 1.75 mm diameter was extruded at 190 degrees C onto a 50 degrees C build plate. Printing used a constant volumetric feed rate (18 mm^3^/min) with variable print-head speed (3-30 mm/s) to generate line widths of 0.1-1.0 mm, corresponding to infill ratios of 10-100% at a fixed line spacing of 1.0 mm. This dynamic infill modulation achieves voxel-specific attenuation as prescribed by the phantom model. The fixed 1.0 mm line spacing is stated explicitly here because it defines a periodic structure within the printed parenchyma at approximately 1 line pair per mm. The completed phantom is shown in Figure 1.

**Figure 1.**
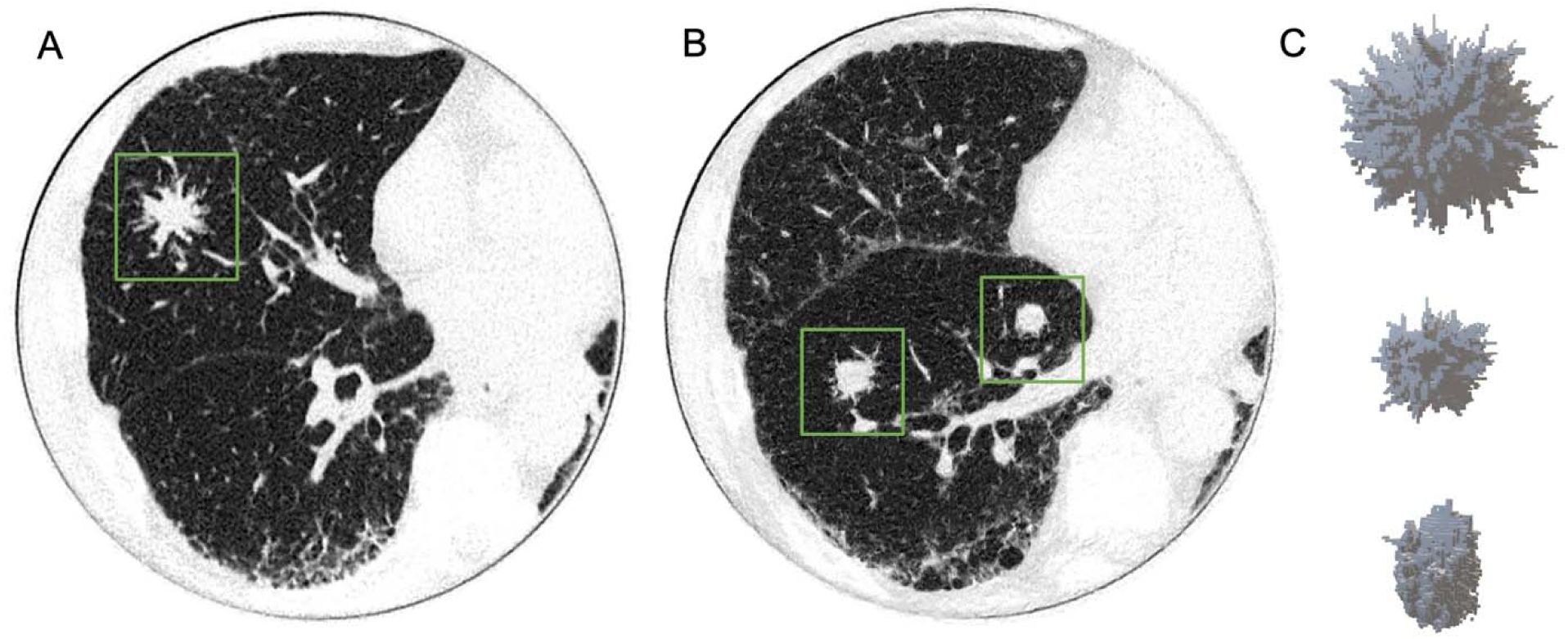
CT images of the 3D-printed ultra-high-resolution lung phantom with inserted digital lesions. Axial views are shown in A and B, with boxes indicating the locations of the inserted lesions. C shows the corresponding 3D lesion models used for phantom generation: a highly spiculated spherical lesion (27 mm), a moderately spiculated spherical lesion (13 mm), and a mildly spiculated ovoid lesion (10 x 17 mm) elongated along the z-axis. Window level/width: −500/1000 HU.

### Phantom Image Acquisition

All phantom acquisitions were performed on a CZT-based photon-counting detector CT prototype (Canon Inc., Otawara, Japan). This system is a research prototype and is not commercially available. The protocol used a tube voltage of 120 kVp and a helical pitch factor of 0.828. Four dose levels were acquired, with volumetric CT dose Index (CTDI_vol_) of 5.8, 3.9, 1.9, and 1.1 mGy (nominal 6, 4, 2, and 1 mGy) at 75, 50, 25, and 14 mAs respectively; gantry rotation time varied correspondingly from 1000 to 350 ms. The printed lung phantom measured approximately 20 cm in diameter and 5 cm in height. To approximate a larger body habitus and modulate x-ray attenuation, it was imaged inside a 3D-printed extension ring (35.5 x 26.5 cm).

### Image Reconstruction

Each acquisition was reconstructed with two algorithms. Hybrid IR (AIDR 3D, Canon Inc.) used the FC52 lung kernel at high strength (level 3) and was generated at both normal resolution (512 x 512) and ultra-high resolution (1024 x 1024). DLR (AiCE, Canon Inc.) used the lung setting at high strength (level 3) and was generated at ultra-high resolution only. Reconstruction diameter was 318 mm, giving a reconstructed pixel size of 0.31 mm, with a slice thickness of 0.21 mm. All reconstructions were performed on the scanner console or affiliated reconstruction platform using vendor-default implementations for the specified settings. Acquisition and reconstruction parameters are summarized in Table 1.

**Table 1.**
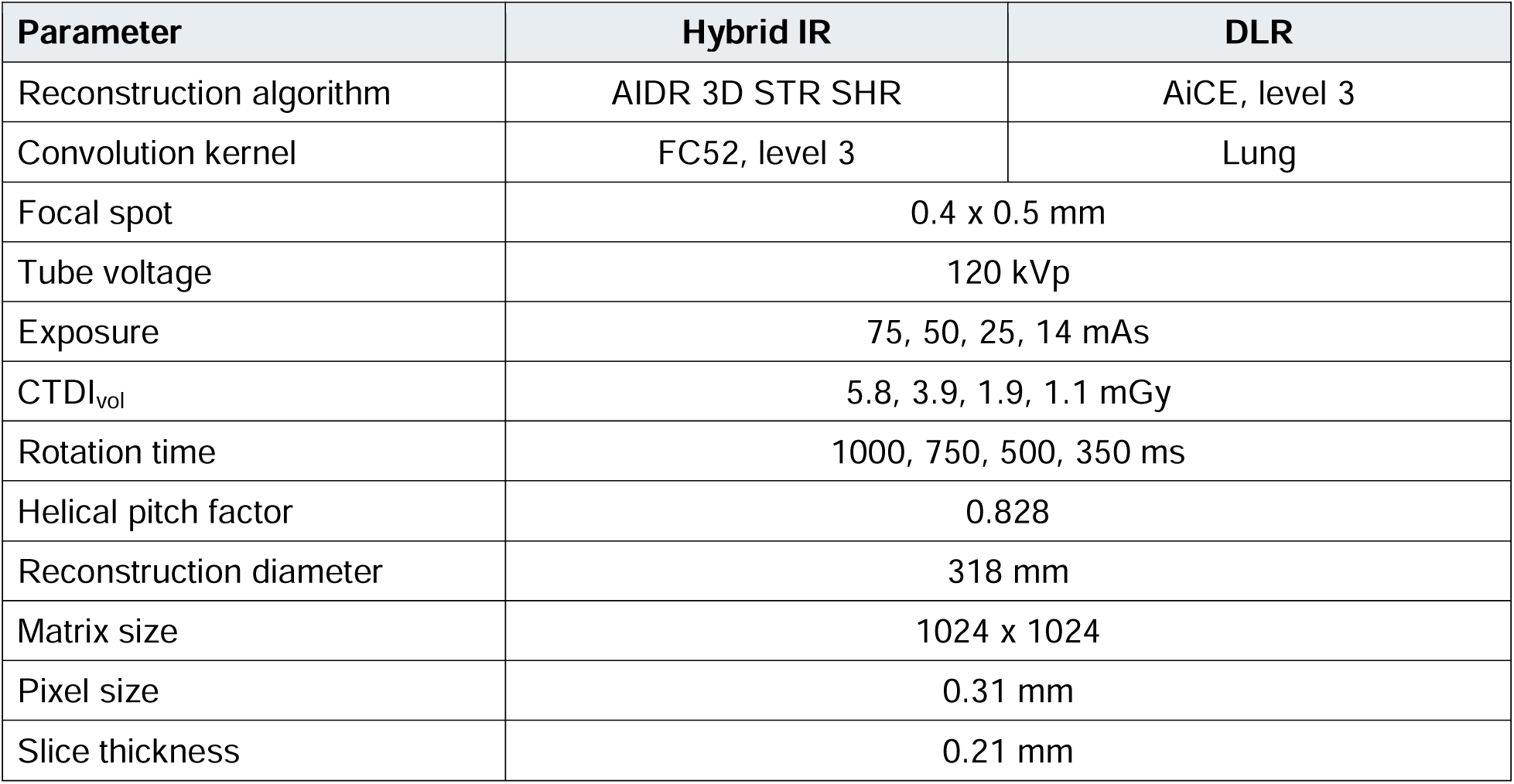
Acquisition and reconstruction parameters, phantom study.

| Parameter | Hybrid IR | DLR |
| --- | --- | --- |
| Reconstruction algorithm | AIDR 3D STR SHR | AiCE, level 3 |
| Convolution kernel | FC52, level 3 | Lung |
| Focal spot | 0.4 x 0.5 mm |  |
| Tube voltage | 120 kVp |  |
| Exposure | 75, 50, 25, 14 mAs |  |
| CTDI<sub>vol</sub> | 5.8, 3.9, 1.9, 1.1 mGy |  |
| Rotation time | 1000, 750, 500, 350 ms |  |
| Helical pitch factor | 0.828 |  |
| Reconstruction diameter | 318 mm |  |
| Matrix size | 1024 x 1024 |  |
| Pixel size | 0.31 mm |  |
| Slice thickness | 0.21 mm |  |

### Image Noise, Lesion Contrast, and Contrast-to-Noise Ratio

Image noise was defined as the standard deviation of attenuation within a region of interest (ROI) placed inside each lesion. Lesion contrast was defined as the difference in mean attenuation between the lesion ROI and an adjacent parenchymal ROI, and contrast-to-noise ratio (CNR) as this contrast divided by the image noise. Contrast is reported separately from CNR because, as shown in the Results, the two are not independent in this dataset. ROI size, shape, and position were prespecified and replicated across reconstructions by coordinate transfer, so that identical anatomical locations were sampled in every image.

### Spatial Resolution

Spatial resolution was assessed using the modulation transfer function (MTF), measured by the slanted-edge method at the interface between the 0 HU and −1000 HU materials within the phantom. Eight separate edge locations were analyzed. Measurements were performed in ImageJ using the Slanted Edge MTF plugin [27]. MTF is reported both as full curves and as MTF50 and MTF10 (the spatial frequencies at which modulation falls to 50% and 10% of its low-frequency value). The edge used spans approximately 1000 HU. Because both reconstruction algorithms are nonlinear, their measured performance depends on the properties of the object being imaged. Metrics obtained under one set of conditions, a given edge contrast, noise level, or structural scale, do not necessarily transfer to others, and this applies to the resolution, noise, and texture measurements reported here.

### Texture Analysis

Local image texture was quantified using gray-level co-occurrence matrices (GLCMs) [28]. A GLCM tabulates how frequently pairs of pixels separated by a defined offset take particular combinations of gray values. Entries concentrated near the main diagonal indicate that neighboring pixels have similar attenuation, consistent with a locally homogeneous appearance; entries displaced from the diagonal indicate greater local variation, consistent with sharper transitions or more complex structure. The extent of the occupied region along the diagonal reflects the range of attenuation values present and is therefore sensitive to partial-volume mixing between materials, whereas displacement perpendicular to the diagonal reflects differences between neighboring pixels. These two properties are reported separately, as they are not interchangeable. GLCM was selected because it is defined identically for phantom and patient images and can therefore be applied to both without modification. GLCM analysis has previously been applied to lung texture [29], [30], [31], [32].

Pixel intensities were discretized into 256 gray levels over a fixed range of −1000 to 0 HU prior to matrix generation for the phantom, and 128 levels over the range of −900 to 0 HU for the patient images. Identical quantization was applied to every image, region, and reconstruction, since GLCM appearance and all derived features depend strongly on both bin count and range. In the phantom, a single ROI was placed within a region of printed parenchyma containing interstitial and vascular structure, measuring 20 x 20 mm^2^ (63 x 63 pixel^2^). The same anatomical region was analyzed across all reconstructions and dose levels by coordinate transfer. Because this region contains both air-filled and tissue-equivalent components rather than uniform parenchyma, attenuation histograms are reported alongside the matrices, and mean attenuation within the region is reported to confirm that attenuation accuracy was preserved across reconstructions.

### Illustrative Patient Examples

Two patient examinations acquired on the same CZT-based PCCT prototype were selected as illustrative examples. Cases were chosen to represent patterns in which fine texture, lesion internal architecture, and margin assessment are relevant to interpretation. Selection was not random, no consecutive series was sampled, and no comparison between cases is made. All patient data were obtained under institutional review board approval (IRB# 24-1434 (856690)), with written informed consent obtained prior to imaging. All data were de-identified prior to analysis. Acquisition and reconstruction parameters followed the departmental thoracic protocol and are summarized in Table 2.

**Table 2.** Acquisition and reconstruction parameters, illustrative patient examples.

| Parameter | Hybrid IR | DLR |
| --- | --- | --- |
| Reconstruction algorithm | AIDR 3D STD SHR | AiCE, level 3 |
| Convolution kernel | FC52 | Lung |
| Focal spot | 0.4 x 0.5 mm |  |
| Tube voltage | 120 kVp |  |
| Exposure | 70 and 52 mAs |  |
| CTDI<sub>vol</sub> | 5.4 and 4.1 mGy |  |
| Rotation time | 350 ms |  |
| Helical pitch factor | 0.828 |  |
| Reconstruction diameter | 438.5 and 403.3 mm |  |
| Matrix size | 1024 x 1024 |  |
| Pixel size | 0.43 and 0.39 mm |  |
| Slice thickness | 0.21 mm |  |

In the patient lesion examples, part-solid lung nodules were selected, with the solid and ground-glass components segmented separately by an experienced radiologist (board-certified chest fellowship-trained with over 13 years of experience), and the segmentations were transferred unchanged to the remaining reconstructions. GLCMs were computed within each segmented component independently, counting only pixel pairs for which both members fell inside the mask.

## Results

### Qualitative Appearance Across Dose and Reconstruction

Representative phantom images comparing UHR hybrid IR and UHR DLR across dose levels are shown in Figure 2. Across the four dose levels, DLR showed reduced visible image noise while fine parenchymal and interstitial structures remained discernible. The difference in visible noise was greater at the lower dose levels, where UHR hybrid IR images showed increased blurring and reduced conspicuity of fine detail.

**Figure 2.**
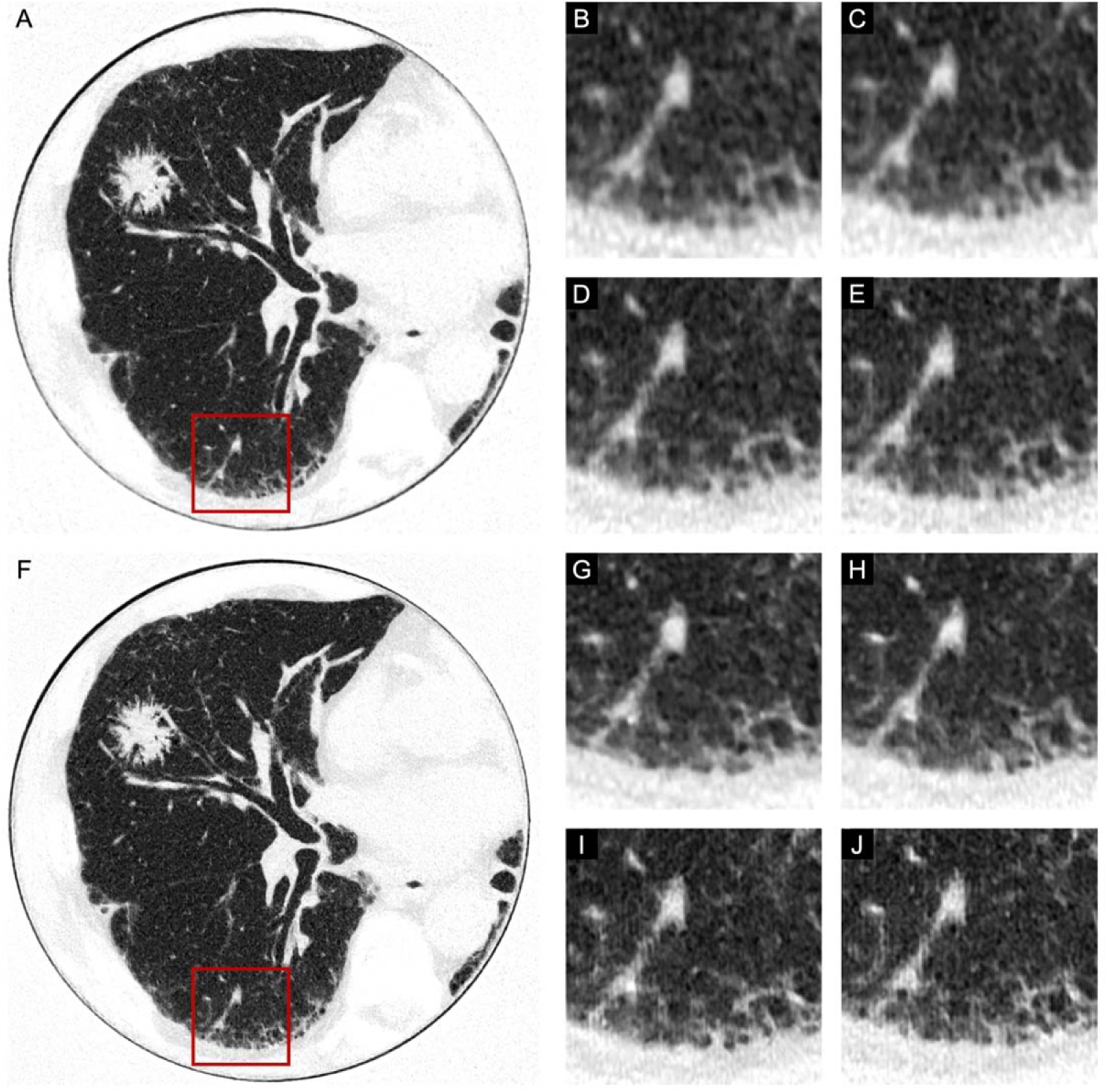
Representative phantom images comparing ultra-high-resolution hybrid IR (A-E) and ultra-high-resolution DLR (F-J) across dose levels. Magnified parenchymal and interstitial regions are shown at CTDI_vol_ 1, 2, 4, and 6 mGy (B-E and G-J). DLR demonstrates reduced visible noise while preserving fine pulmonary structure across dose levels. Window level/width: −500/1000 HU.

Magnified views centered on the three inserted lesions are shown in Figure 3. At both 6 and 1 mGy, DLR showed lower apparent image noise and more clearly delineated lesion margins than UHR hybrid IR. This was most evident for the spiculated component, where fine peripheral projections were better separated from the background.

**Figure 3.**
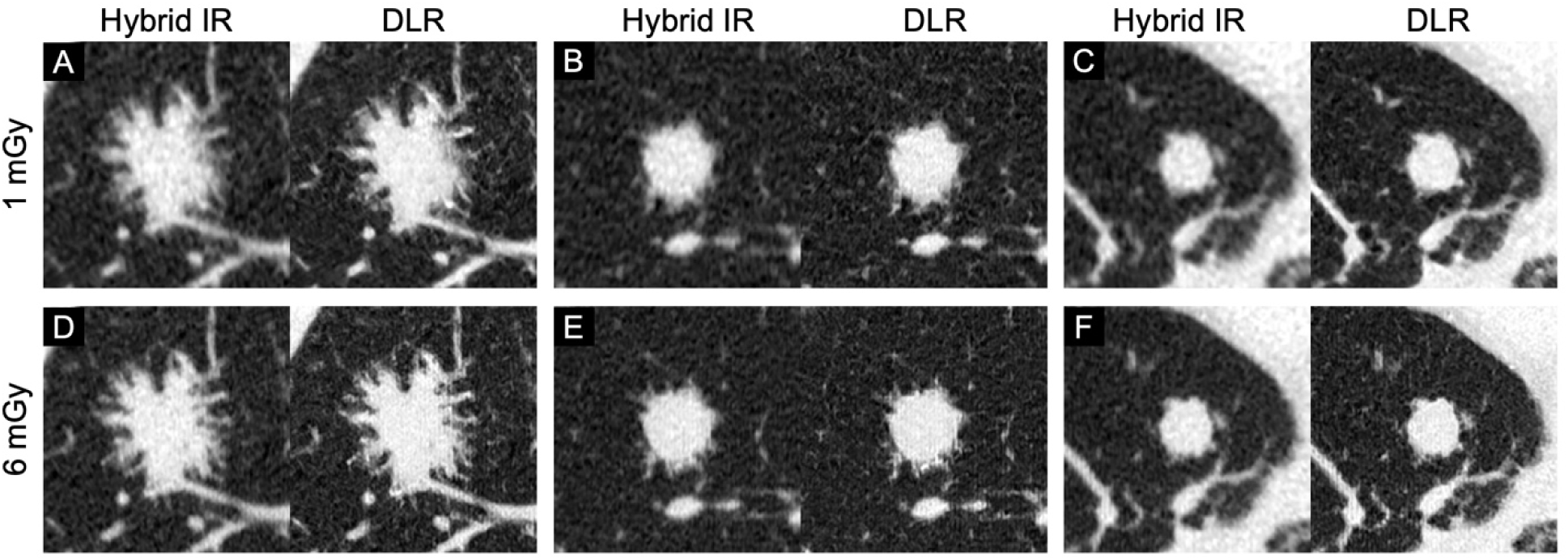
Magnified views centered on Lesions 1, 2, and 3 (columns) at CTDI_vol_ 1 mGy (upper row) and 6 mGy (lower row). For each lesion, paired images compare ultra-high-resolution hybrid IR and ultra-high-resolution DLR. Window level/width: −500/1000 HU.

### Image Noise, Lesion Contrast, and Contrast-to-Noise Ratio

At 6 mGy, image noise within lesion ROIs was lower with DLR than with UHR hybrid IR for Lesion 1 (32.9 ± 1.2 vs 36.9 ± 1.4 HU; 10.9% lower) and Lesion 2 (34.2 ± 2.3 vs 39.1 ± 1.7 HU; 12.5% lower), and was unchanged for Lesion 3 (36.7 ± 1.8 vs 36.6 ± 2.6 HU). At 1.1 mGy, noise was lower with DLR for Lesion 1 (32.1 ± 4.0 vs 40.7 ± 2.6 HU; 21.1% lower) and Lesion 3 (25.7 ± 0.6 vs 34.1 ± 1.1 HU; 24.6% lower), and was unchanged for Lesion 2 (36.5 ± 1.9 vs 36.5 ± 0.8 HU). On average, this is about −7.8% at 6 mGy and −15.2% at 1 mGy. Lesion contrast, calculated as the difference in mean attenuation between lesion and adjacent parenchymal ROIs, was effectively constant across reconstruction methods and dose levels (measurements in Table 3). Because contrast did not vary, differences in CNR in this dataset are determined entirely by differences in measured noise. CNR values are reported in Table 3 for completeness.

**Table 3.** Image noise, lesion contrast, and contrast-to-noise ratio.

| Dose | Metric | Lesion 1<br>Hybrid IR | Lesion 1<br>DLR | Lesion 2<br>Hybrid IR | Lesion 2<br>DLR | Lesion 3<br>Hybrid IR | Lesion 3<br>DLR |
| --- | --- | --- | --- | --- | --- | --- | --- |

| Dose | Metric | Lesion 1 Hybrid IR | Lesion 1 DLR | Lesion 2 Hybrid IR | Lesion 2 DLR | Lesion 3 Hybrid IR | Lesion 3 DLR |
| --- | --- | --- | --- | --- | --- | --- | --- |
| 6 mGy | Noise (HU) | 36.9 ± 1.4 | 32.9 ± 1.2 | 39.1 ± 1.7 | 34.2 ± 2.3 | 36.6 ± 2.6 | 36.7 ± 1.8 |
|  | Contrast (HU) | 785.7 | 785.6 | 774.4 | 770.7 | 781.5 | 782.4 |
|  | CNR | 21.3 | 23.9 | 19.8 | 22.5 | 21.3 | 21.3 |
| 4 mGy | Noise (HU) | 38.0 ± 2.7 | 32.9 ± 1.0 | 37.7 ± 0.8 | 34.2 ± 1.3 | 44.2 ± 1.1 | 36.7 ± 2.0 |
|  | Contrast (HU) | 777.2 | 782.8 | 776.2 | 778.0 | 783.5 | 786.0 |
|  | CNR | 20.4 | 20.8 | 20.6 | 20.6 | 17.7 | 17.7 |
| 2 mGy | Noise (HU) | 34.3 ± 0.7 | 30.1 ± 3.7 | 31.9 ± 1.5 | 31.5 ± 2.8 | 29.1 ± 2.3 | 27.0 ± 4.7 |
|  | Contrast (HU) | 780.8 | 791.5 | 779.2 | 786.1 | 780.6 | 790.4 |
|  | CNR | 22.8 | 26.3 | 24.4 | 25.0 | 26.9 | 29.3 |
| 1 mGy | Noise (HU) | 40.7 ± 2.6 | 32.1 ± 4.0 | 36.5 ± 0.8 | 36.5 ± 1.9 | 34.1 ± 1.1 | 25.7 ± 0.6 |
|  | Contrast (HU) | 794.8 | 794.0 | 780.0 | 787.1 | 781.4 | 791.3 |
|  | CNR | 19.5 | 24.7 | 21.3 | 21.5 | 22.9 | 30.8 |

### Spatial Resolution

Spatial resolution results are shown in Figure 4 as MTF plotted against spatial frequency (lp/mm) for hybrid IR and DLR at CTDI_vol_ 6 mGy and 1 mGy. At both dose levels, the DLR curves remained above the corresponding hybrid IR curves across the displayed spatial frequency range, indicating improved preservation of higher-frequency image information. This difference was observed at both the higher and lower dose settings, suggesting that DLR maintained spatial resolution advantages even under increased noise conditions. MTF50 for hybrid IR6 and 1 mGy are 0.8 and 0.6, for DLR 6 and 1 mGy are 1.2 and 1.0. MTF10 for hybrid IR 6 and 1 mGy are 2.2 and 1.9, for DLR 6 and 1 mGy are 2.6 and 2.5, estimated by linear interpolation.

**Figure 4.**
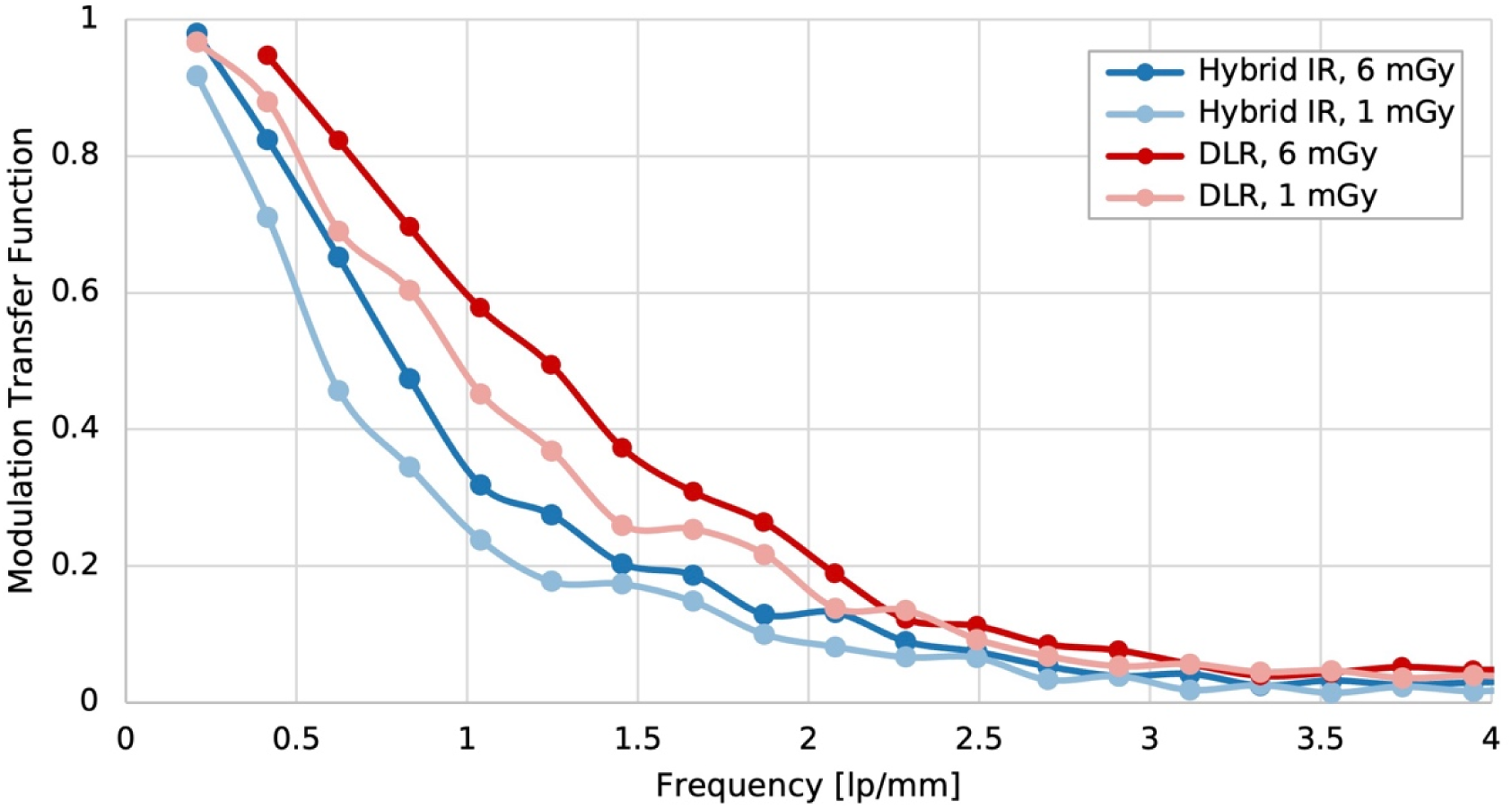
Modulation transfer function (MTF) curves for ultra-high-resolution hybrid IR and DLR at CTDI_vol_ 6 mGy and 1 mGy. DLR demonstrated higher MTF values across the displayed spatial frequency range at both dose levels, consistent with improved preservation of high-frequency image information.

### Texture Analysis in the Phantom

Texture analysis of the phantom region is shown in Figure 5 for hybrid IR and DLR at 1 and 6 mGy. The selected region contains a network of septal and vascular structures against lower-attenuation parenchyma, and the corresponding histograms are correspondingly broad and right-skewed, with a mode near −820.3 HU and a tail extending to approximately −321.8 HU. Histogram shape differed systematically across reconstructions. The mode became progressively narrower from hybrid IR at 1 mGy through to DLR at 6 mGy, 319, 222, 208, 206 HU, while the position of the mode and the extent of the high-attenuation tail were preserved. Because the mode corresponds to the air-filled component of the region, this narrowing, about 23%, is consistent with reduced mixing between air and tissue attenuation values, that is, with reduced partial-volume averaging and noise, rather than with a shift in attenuation. Mean attenuation within the region was −648.7 HU and varied by less than 2.2 HU across all four reconstructions, indicating that attenuation accuracy was maintained. The corresponding GLCMs are shown in the bottom row. GLCM for DLR at 6 mGy showed the broadest distribution away from the main diagonal, consistent with greater local intensity variation and more complex texture.

**Figure 5.**
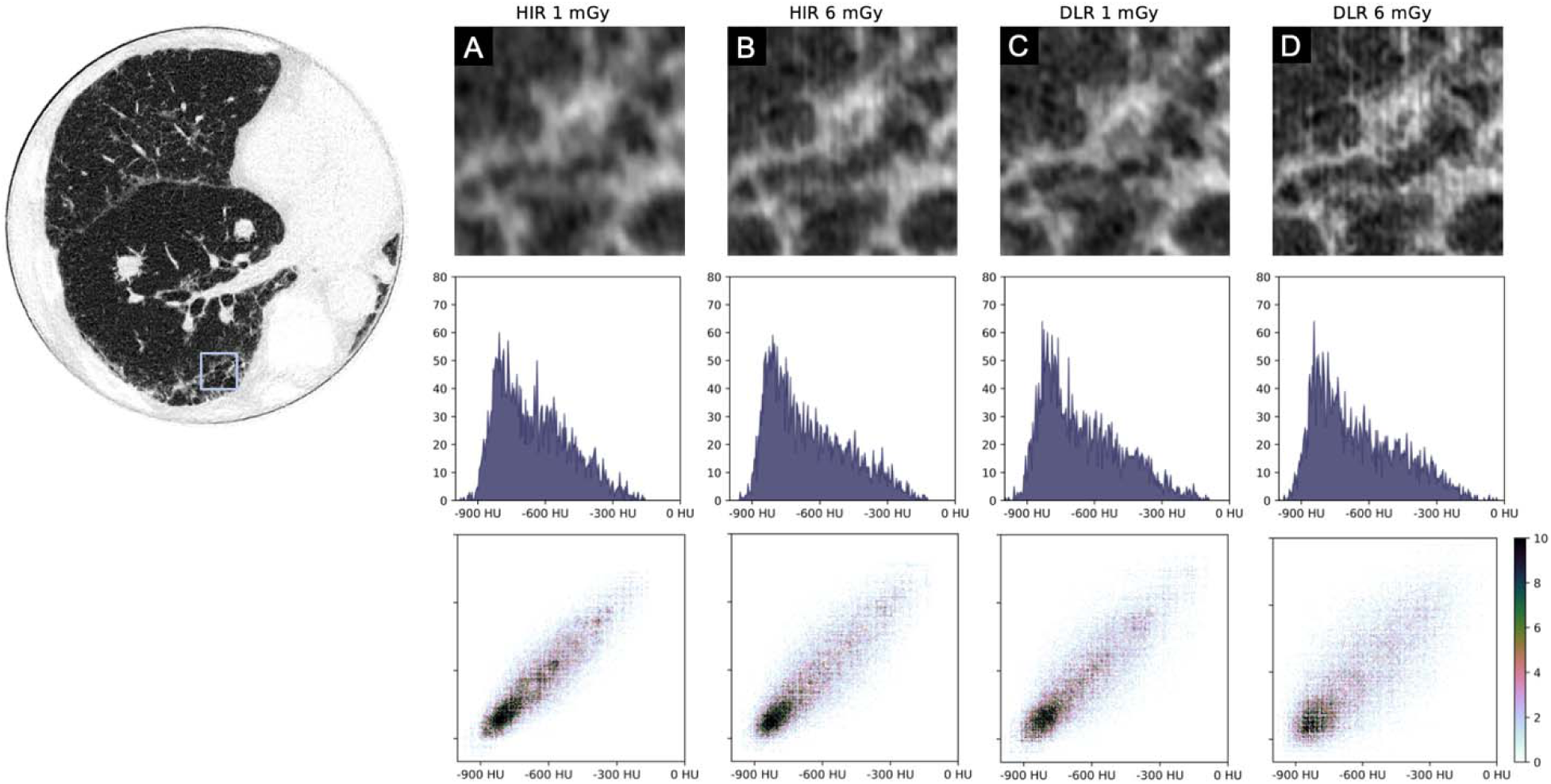
Texture analysis of an interstitial region in the printed lung phantom for hybrid IR (HIR) and DLR at CTDI_vol_ 1 mGy and 6 mGy. The top row shows the selected regions of interest for (A) hybrid IR at 1 mGy, (B) hybrid IR at 6 mGy, (C) DLR at 1 mGy, and (D) DLR at 6 mGy. The middle row shows the corresponding histograms of pixel intensities from −1000 HU to 0 HU. The bottom row shows the corresponding GLCMs. GLCM for DLR at 6 mGy showed the broadest distribution away from the main diagonal, consistent with greater local intensity variation and more complex texture. In addition, fine printing structures became visible in the DLR 6 mGy image, further illustrating the ability of DLR to preserve or recover subtle high-resolution detail.

### Illustrative Patient Examples

#### Example 1: Part-Solid Lesion

A female patient in her 50’s with a part-solid pulmonary nodule in the right lower lobe is shown in Figure 6. On normal-resolution hybrid IR, the lesion appeared as an ill-defined nodule with indistinct margins and no discernible internal architecture. Ultra-high-resolution hybrid IR better resolved internal structure and the adjacent vessel, as well as a small, poorly delineated solid component at the upper medial aspect of the lesion, with a corresponding increase in visible image noise. Ultra-high-resolution DLR showed the same structures with lower apparent noise, as well as a better visualization of the vessel entering the lesion inferiorly, and the boundary between the ground-glass and solid components was more distinctly separated from surrounding parenchyma and from one another.

**Figure 6.**
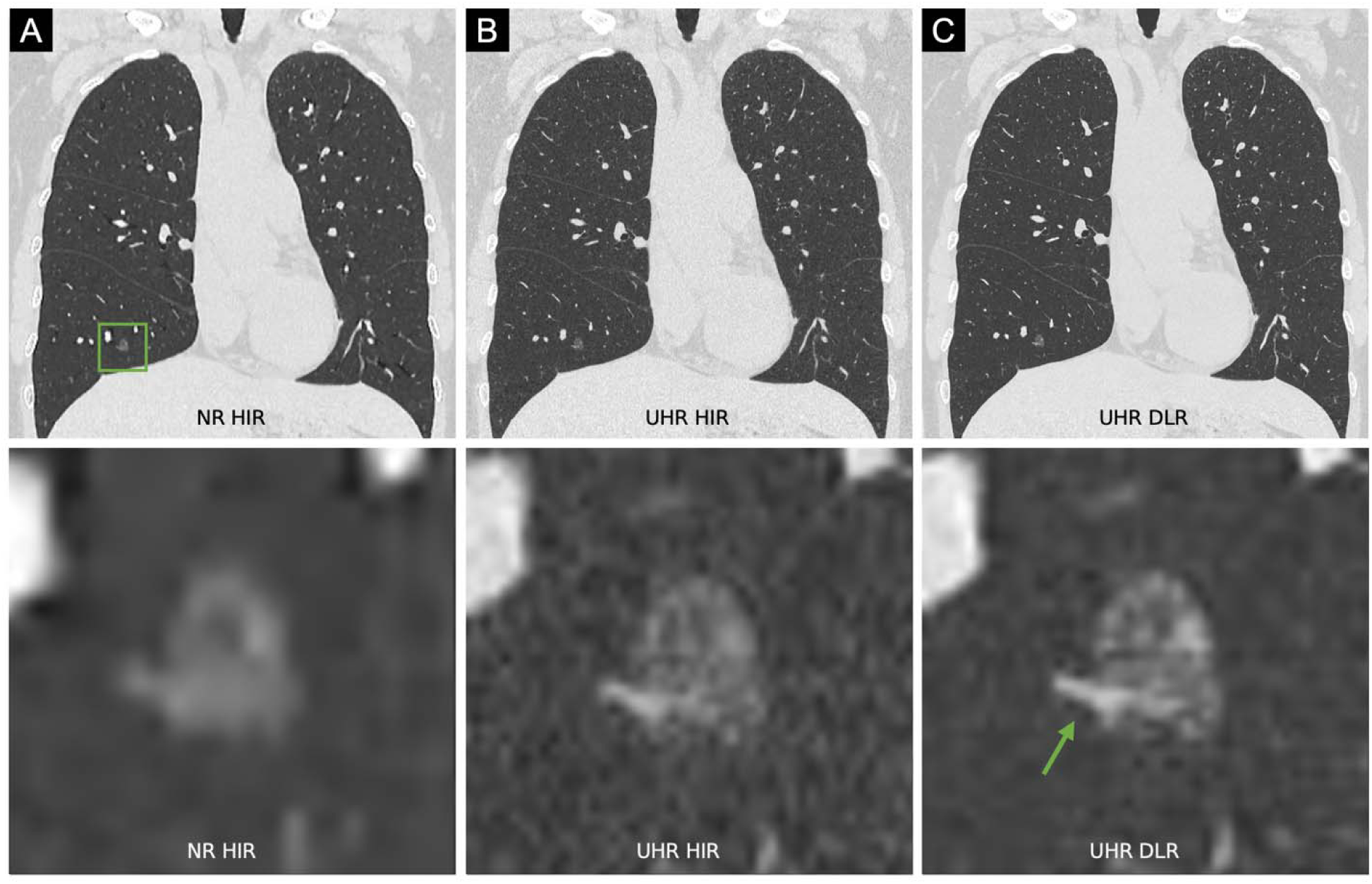
Illustrative patient example 1: part-solid lesion in the right lower lobe. Upper row: coronal images reconstructed with (A) normal-resolution (NR) hybrid IR (HIR), (B) ultra-high-resolution (UHR) hybrid IR, and (C) ultra-high-resolution DLR. The green box in (A) indicates the location of the lesion. Lower row: magnified views of the boxed region for the corresponding reconstructions. Window level/width: −550/1600 HU.

To characterize this observation quantitatively, the solid and ground-glass components were segmented separately and analyzed with the same operator used in the phantom (Figure 7). Attenuation histograms of the two components overlapped substantially on normal-resolution hybrid IR by 6.3% and on ultra-high-resolution hybrid IR by 7.7%, with separability of 1.59 and 1.62. The ground-glass component centered at −675.0 and −709.5 HU respectively and the solid component at −505.5 and −553.0 HU. For ultra-high-resolution DLR, the two distributions were more isolated compared to either IR, with overlapped by 5.1% and a higher separability of 2.04. The ground-glass component centered at about same attenuation (−678.0 HU) but the solid component at a higher attenuation (−392.0 HU), compared to either hybrid IR. The direction of these shifts is consistent with reduced partial-volume mixing between the two components: with sharper rendering, voxels within each component are less influenced by the attenuation of the adjacent one, so each distribution moves toward its own characteristic value rather than toward the other. Gray-level co-occurrence matrices computed within each segmented component are shown in the lower row of Figure 7. The matrices for the two components occupied distinct positions along the main diagonal, and this separation was greatest on ultra-high-resolution DLR, mirroring the histogram findings. Because the segmented components in this example contain 218 and 54 pixels, the corresponding matrices are sparsely populated and are presented as illustration.

**Figure 7.**
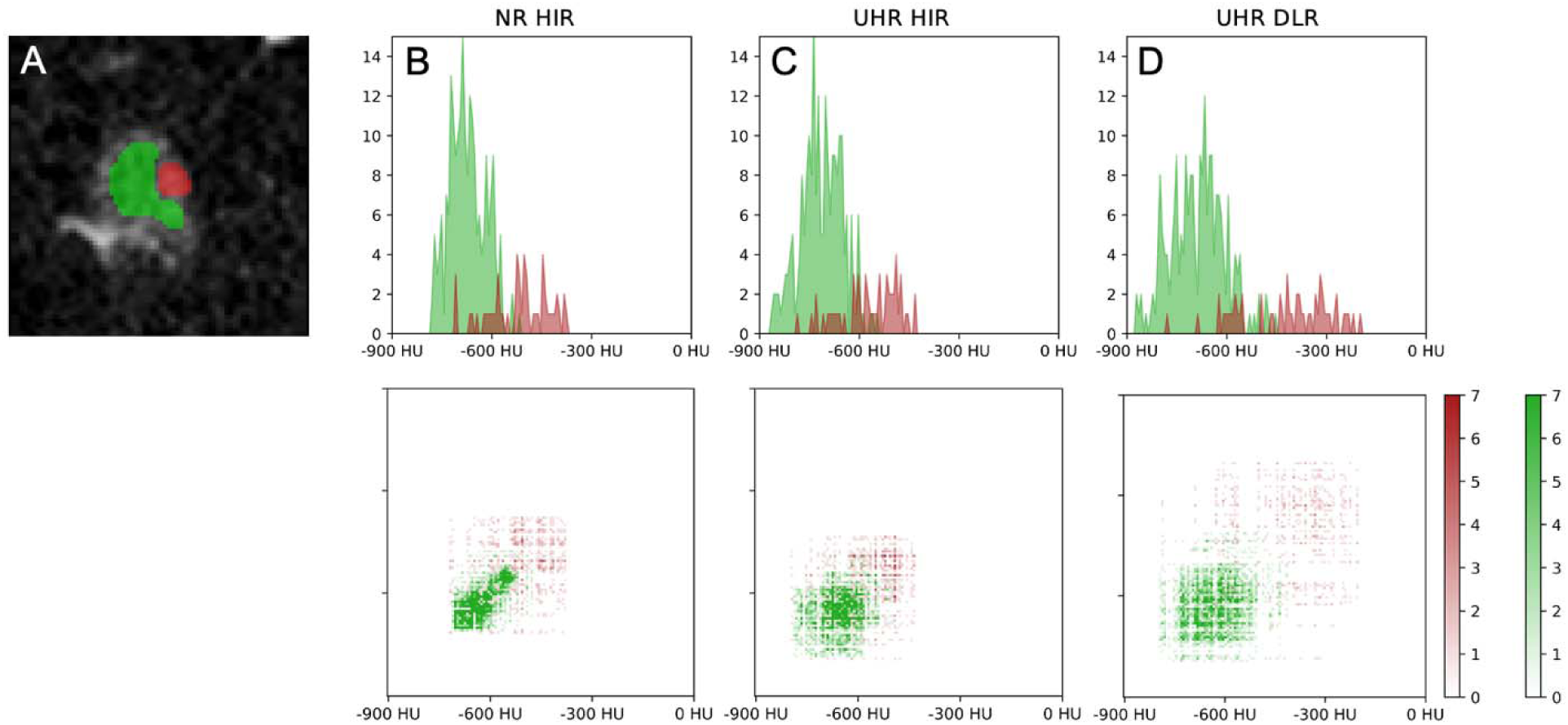
Component analysis of the lesion shown in Figure 6. Upper row: (A) Magnified view of the ultra-high-resolution (UHR) DLR lesion with the segmented solid component (red) and ground-glass component (green). (B–D) Attenuation histograms of the two segmented components for (B) normal-resolution hybrid IR (HIR), (C) ultra-high-resolution hybrid IR, and (D) ultra-high-resolution DLR. The vertical axis indicates pixel counts. Lower row: gray-level co-occurrence matrices for the corresponding segmentations and reconstructions, with color bars indicating counts.

#### Example 2: Part-Solid Lesion with Linear Architecture

A female patient in her late 70’s with a part-solid lesion in the right lower lobe is shown in Figure 8. The lesion had an elongated configuration with an internal linear structure of higher attenuation extending along its long axis.

**Figure 8.**
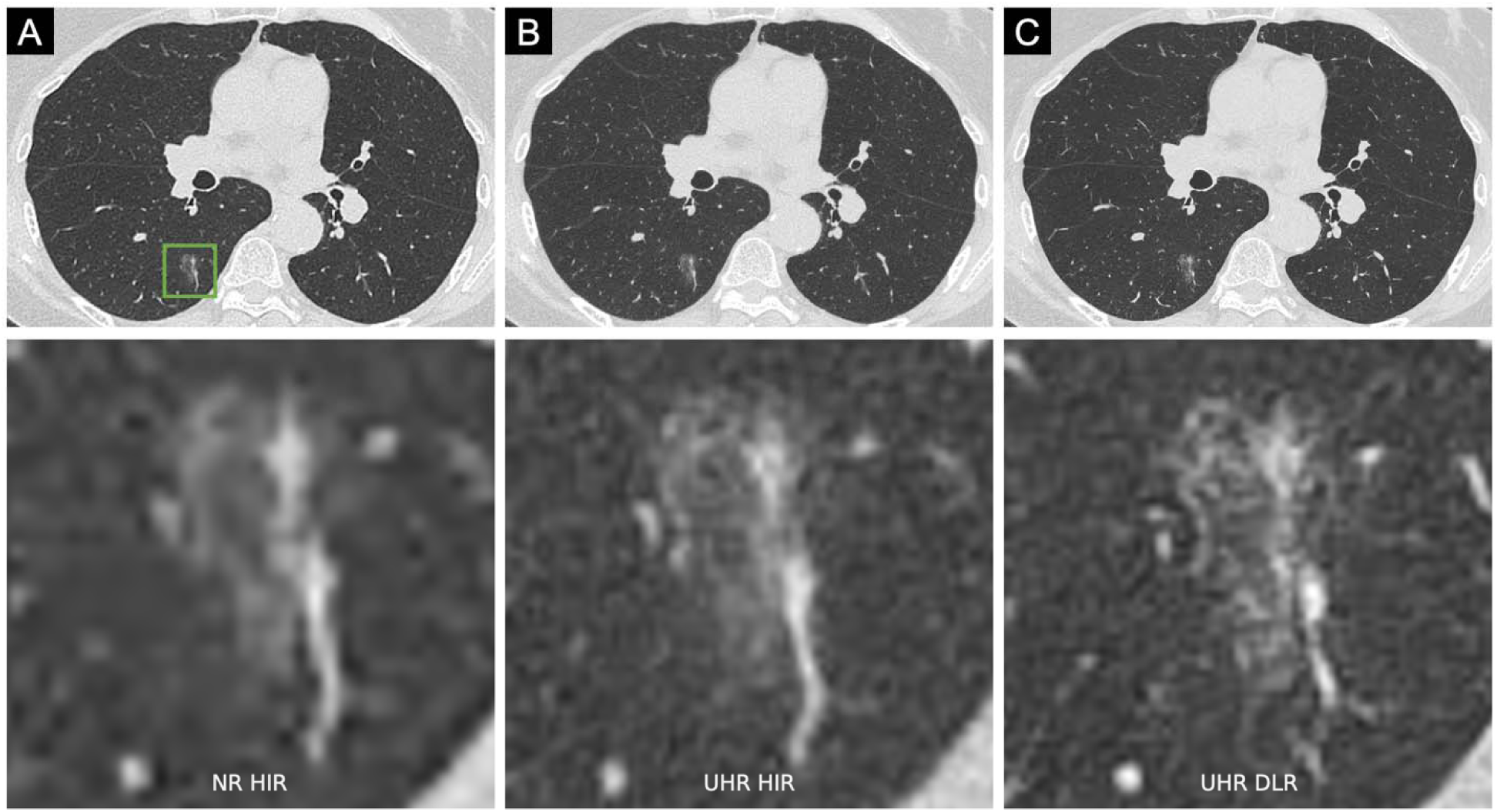
Illustrative patient example 2: part-solid lesion with internal linear architecture in the right lower lobe. Upper row: axial images reconstructed with (A) normal-resolution (NR) hybrid IR (HIR), (B) ultra-high-resolution hybrid IR, and (C) ultra-high-resolution deep DLR. The green box in (A) indicates the location of the lesion. Lower row: magnified views of the boxed region for the corresponding reconstructions. Window level/width: −550/1600 HU.

On normal-resolution hybrid IR, the lesion appeared as an ill-marginated focus with the internal linear structure blurred and its boundary with the surrounding ground-glass component was indistinct. Ultra-high-resolution hybrid IR resolved the linear structure and adjacent small vessels, with increased visible image noise. UHR DLR showed the same structures with lower apparent noise, and the interface between the linear component and the surrounding ground-glass component was more distinctly delineated.

Solid and ground-glass components were segmented separately and analyzed with the operator used for the phantom and for patient example 1 (Figure 9). On all three reconstructions, the two components occupied distinct but partially overlapping attenuation ranges, with the ground-glass component centered near −684.2 HU and the solid component distributed above approximately −328.0 HU. Two changes were apparent across reconstructions. First, the solid component distribution shifted toward higher attenuation, from median −312.5 HU on normal-resolution hybrid IR and −322.0 HU on ultra-high-resolution hybrid IR and −296.5 HU on ultra-high-resolution DLR. Second, the ground-glass distribution became wider on ultra-high-resolution DLR, extending to lower attenuation values than on either hybrid IR. Gray-level co-occurrence matrices for the two segmented components are shown in the lower row of Figure 9.

**Figure 9.**
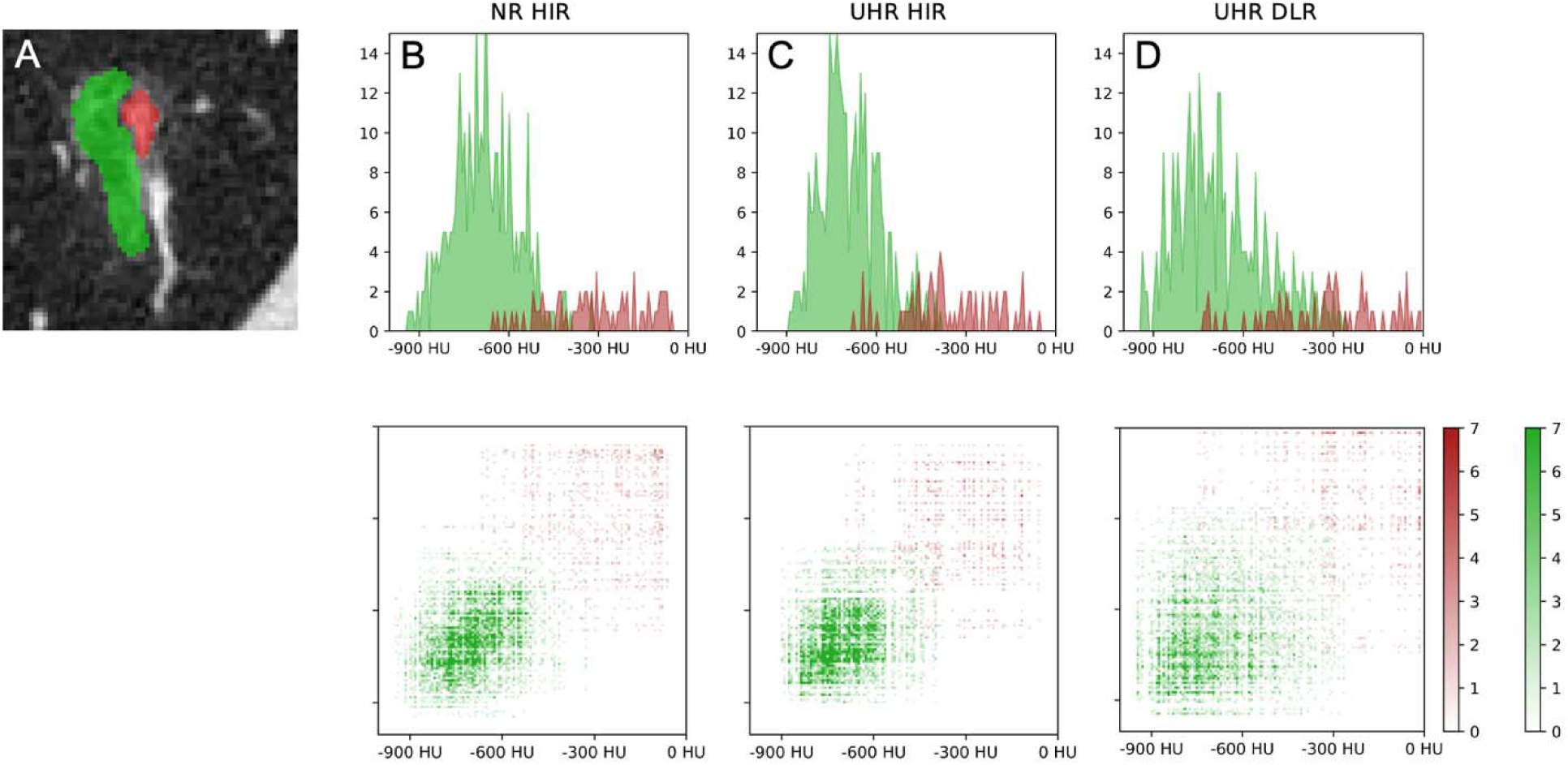
Component analysis of the lesion shown in Figure 8. Upper row: (A) Magnified view with the segmented solid component (red) and ground-glass component (green). (B–D) Attenuation histograms of the two segmented components for (B) normal-resolution (NR) hybrid IR (HIR), (C) ultra-high-resolution (UHR) hybrid IR, and (D) ultra-high-resolution DLR. The vertical axis indicates pixel counts. Lower row: gray-level co-occurrence matrices for the corresponding segmentations and reconstructions, with separate color bars for the two components indicating counts.

The matrix for the ground-glass component extended over a wider range of attenuation values on ultra-high-resolution DLR than on either hybrid IR, consistent with the histogram findings.

The segmented components contain 340 and 70 pixels, so the matrices are sparsely populated relative to the number of matrix cells and are presented as an illustration.

## Discussion

We characterized DLR against hybrid IR for ultra-high-resolution photon-counting CT of the lung, using a patient-derived 3D-printed phantom, and applied the same analytical framework to two patient examinations acquired on the same system. In the phantom, DLR reduced image noise relative to UHR hybrid IR. Lesion contrast was preserved across all reconstructions and dose levels, so the corresponding CNR differences reflect the change in noise alone rather than an independent change in lesion conspicuity. MTF curves for DLR lay above those for hybrid IR across the evaluated frequency range at both dose levels. Attenuation histograms of a structured parenchymal region showed a progressively taller and narrower mode from hybrid IR at 1 mGy through DLR at 6 mGy, with the mode position and the high-attenuation tail preserved and mean attenuation stable. In both patient examples, the attenuation distributions of the segmented solid and ground-glass components were more widely separated on UHR DLR than on either hybrid IR.

The consistent thread across these observations is a reduction in partial-volume mixing rather than a change in attenuation. A voxel spanning two materials reports their average, so blurring pulls air-filled voxels upward and tissue voxels downward, compressing the distance between the two populations while leaving their combined mean unchanged. Sharper rendering reverses that compression. This predicts precisely what was observed: a narrower air-end mode in the phantom with the mode position unmoved, a stable regional mean, and greater separation between solid and ground-glass components in the patient examples without a shift in either component’s characteristic value. The same mechanism accounts for the qualitative impression of better-delineated lesion margins, since a margin is an interface between two attenuation populations and its apparent sharpness is the degree to which they remain distinct.

The contribution of the phantom is not that it permits a comparison of two reconstruction algorithms, which has been reported on this platform in patients [19], but that it constrains how such a comparison can be read with regard to different radiation dose levels. In patient images, a reconstruction that renders parenchyma with more apparent fine detail cannot be distinguished from one that generates it, because the ground truth is unavailable. A patient-derived printed phantom supplies a partial reference, since the prescribed attenuation of every voxel is known and the source dataset from which the phantom was printed represents the texture the phantom is intended to reproduce. In the present work this reference was used to confirm that regional attenuation was preserved across reconstructions, which is what connects to the partial-volume interpretation above.

Gray-level co-occurrence matrices provide a compact visual summary of how attenuation values are jointly distributed among neighboring pixels, and because the descriptor is defined identically for phantom and patient images, it can be applied on both sides without modification. In the phantom, the matrices show the range of attenuation values present within the analyzed region and how tightly neighboring pixels are coupled across it; in the patient examples, the matrices for the solid and ground-glass components occupy distinct positions along the main diagonal, so the separation between components is directly visible in a single representation. This makes the matrices a useful complement to the histogram measurements, displaying jointly what the histograms describe marginally. Their interpretive scope is correspondingly specific. The matrices are presented here in visual form and were not reduced to scalar texture features, so the quantitative comparisons in this study rest on the noise, contrast, and attenuation-distribution measurements, with the co-occurrence matrices serving to display the same effects in a spatially explicit form. This is also the appropriate reading given the size of the segmented patient components, which are small relative to the number of matrix elements. A broader co-occurrence distribution is likewise consistent with several underlying causes, preserved anatomical structure, residual noise and edge enhancement.

Several limitations bear directly on how these results should be interpreted. First, the ultra-high-resolution mode operated at the limits of the phantom fabrication process. The printed parenchyma contains a periodic structure, which is resolvable at the reconstructed pixel size and became visible in the higher-dose UHR images. This has two consequences that pull in opposite directions. It demonstrates that the combination of UHR acquisition and DLR resolves physical structure at the fabrication scale, which is itself informative. But it also means that texture measurements made in printed parenchyma at these settings contain a contribution from the printing pattern that was not separately quantified in this study, and the co-occurrence findings in that condition should be interpreted with this in view. Second, the patient component consists of two examinations, selected rather than sampled, and both are part-solid lesions. These cases demonstrate that the analytical framework defined in the phantom can be applied to clinical data and that the direction of the component-separation effect is the same in both. They do not establish that the observed image-quality differences translate into improved diagnostic accuracy, reader confidence, or clinical decision-making, and no such claim is made. Finally, the scanner is a research prototype. Detector configuration, correction chains, and reconstruction implementations may differ from any commercially released system, and results should not be assumed to transfer.

In the future, larger clinical studies are needed to determine whether the differences described here affect performance on specific thoracic tasks, particularly quantification of the solid component of subsolid nodules, where the density-separation effect observed in both patient examples has an obvious point of application. Multi-reader studies would establish the relationship between these technical metrics and diagnostic confidence.

## Conclusion

In a patient-derived 3D-printed lung phantom imaged on a CZT-based photon-counting CT prototype, DLR reduced image noise relative to ultra-high-resolution hybrid IR while preserving lesion contrast and regional attenuation and yielded higher MTF values at a high-contrast edge. Attenuation distributions were consistent with reduced partial-volume mixing at material boundaries rather than with a change in attenuation accuracy. In two patient examinations analyzed with the same framework, the attenuation distributions of solid and ground-glass lesion components were more widely separated with DLR than with either hybrid IR, in the same direction as the phantom findings. By characterizing DLR against a phantom with prescribed attenuation, these results provide a physical basis for the improved depiction of fine pulmonary structure reported in patients, and define the specific effect, improved separation of adjacent attenuation components, that clinical evaluation should now test.

## Availability of data and materials

The datasets generated and analysed during the current study are available from the corresponding author on reasonable request.

## Acknowledgements

We acknowledge support through the National Institutes of Health (R01EB035092, R01EB035908) and Canon Inc..

## Conflict of Interest

P. B. Noel has a research agreement with Canon Inc. S. Sharma, S. Ross and Z. Yu are employees of Canon Medical Research USA. S. S. Haliburton and R. Thompson are employees of Canon Healthcare USA. N. Akino and T. Stroud are employees of Canon Inc. Japan.

## Notes

### Competing Interest Statement

The authors have declared no competing interest.

### Author Declarations

All patient data were obtained under institutional review board approval (IRB# 24-1434 (856690)), with written informed consent obtained prior to imaging. All data were de-identified prior to analysis.

